# Prevalence and impact of *oprD* mutations in *Pseudomonas aeruginosa* strains in cystic fibrosis

**DOI:** 10.1101/19013870

**Authors:** Laura J. Sherrard, Bryan A. Wee, Christine Duplancic, Kay A. Ramsay, Keyur A. Dave, Emma Ballard, Claire E. Wainwright, Keith Grimwood, Hanna E. Sidjabat, David M. Whiley, Scott A. Beatson, Timothy J. Kidd, Scott C. Bell

## Abstract

Defective OprD porins contribute to carbapenem resistance and may be important in *Pseudomonas aeruginosa* adaptation to cystic fibrosis airways. It is unclear whether *oprD* mutations are fixed in populations of shared strains that are transmitted between patients or whether novel variants arise during infection. We investigated *oprD* sequences and antimicrobial resistance of two common Australian shared strains, constructed *P. aeruginosa* mutants with the most common *oprD* allelic variants and compared characteristics between patients with or without evidence of infection with strains harbouring these variants. Our data show that three independently acquired nonsense mutations arising from a 1-base pair substitution are fixed in strain sub-lineages. These nonsense mutations are likely to contribute to reduced carbapenem susceptibility in the sub-lineages without compromising *in vitro* fitness. Not only was lung function worse among patients infected with strains harbouring the nonsense mutations than those without, but they also had an increased hazard rate of lung transplantation/death. Our findings further highlight that understanding adaptive changes may help to distinguish patients with greater adverse outcomes despite infection with the same strain.

## INTRODUCTION

Cystic fibrosis (CF) is a life-shortening autosomal recessive disease resulting in progressive lung damage (Elborn, Ramsey et al., 2016). Lower airway infection with *Pseudomonas aeruginosa* is common, increases with age and, when chronic infection is established, is associated with increased antimicrobial resistance (AMR) (Murray, Egan et al., 2007). Although diverse *P. aeruginosa* genotypes are ubiquitous in the natural environment, some people with CF are infected with clonally-related strains (“shared stains”), which often display AMR to multiple drugs (Parkins, Somayaji et al., 2018). Shared strains have been identified nationally in Australia with three genotypes prominent: AUST-01, AUST-02 and AUST-06 (the latter two are particularly common in the state of Queensland, Australia) (Kidd, Ramsay et al., 2013).

Adaptation of *P. aeruginosa* to survive and persist in the CF lower airway microenvironment occurs in the setting of long-term antibiotic selective pressure with the occurrence of substantial intra-strain phenotypical and genotypical population diversity. Genome sequencing studies have demonstrated that certain loci are more prone to mutation than others (e.g. *mexZ, gyrA*) (Dettman, Rodrigue et al., 2013, Diaz Caballero, Clark et al., 2015, Marvig, Dolce et al., 2015a, Marvig, Johansen et al., 2013, Marvig, Sommer et al., 2015b, Mowat, Paterson et al., 2011, Smith, Buckley et al., 2006, Williams, Evans et al., 2015). However, there is limited information on whether mutations in AMR-related loci are fixed within independent shared strain genotypes or whether novel variants are generally observed in individual patient isolates, suggesting parallel evolution. Identifying differences in adaptive changes in *P. aeruginosa* may help to predict people with CF at the greatest risk of lung function deterioration (Hoffman, Kulasekara et al., 2009, Li, Kosorok et al., 2005, Mayer-Hamblett, Rosenfeld et al., 2014, Tai, Bell et al., 2015a).

We have performed a detailed analysis of *oprD* encoding an outer membrane porin, OprD (Porin D or Protein D2 [(Winsor, Griffiths et al., 2016)]), and AMR in a longitudinal collection of shared strains from patients with CF. Polymorphisms in *oprD* have been detected frequently within, and between, strains in many studies indicating their potential importance in adaptation of *P. aeruginosa* to the CF airways (Greipel, Fischer et al., 2016, Kos, Deraspe et al., 2015, Marvig et al., 2013, Marvig et al., 2015b, Wee, Tai et al., 2018). OprD facilitates diffusion of substrates, such as carbapenems and basic amino acids, across cell membranes. It is well recognised that decreased expression or mutational inactivation of *oprD* is an important AMR mechanism (Courtois, Caspar et al., 2018, Jaillard, van Belkum et al., 2017, Kos et al., 2015, Strateva & Yordanov, 2009). It has been shown that virulence is enhanced in *P. aeruginosa* lacking OprD (Skurnik, Roux et al., 2013), demonstrating that mutations at this locus could potentially have consequences for *in vivo* pathogenicity.

In this study we hypothesised that commonly encountered shared *P. aeruginosa* strains derived from patients with CF harbour loss-of-function *oprD* mutations that: (1) contribute to reduced susceptibility to carbapenems, and (2) could be used as genetic markers to identify a subset of patients at higher risk of an adverse clinical course.

## RESULTS

### *In silico* analysis of *oprD* sequences

The phylogenetic relationship of *oprD* variants is shown in Fig 1A. The wild-type variant of AUST-02 and AUST-06 (1326-bp) is denoted as the variant that is closest to the base of each *oprD* clade. These are highly conserved with only 2 nucleotide differences between the lineages at nucleotide positions 807 (T or C) and 1065 (C or G); their protein sequences are identical (441 amino acids). Fourteen of 114 (12%) isolates had the denoted wild-type variant.

**Fig 1.**
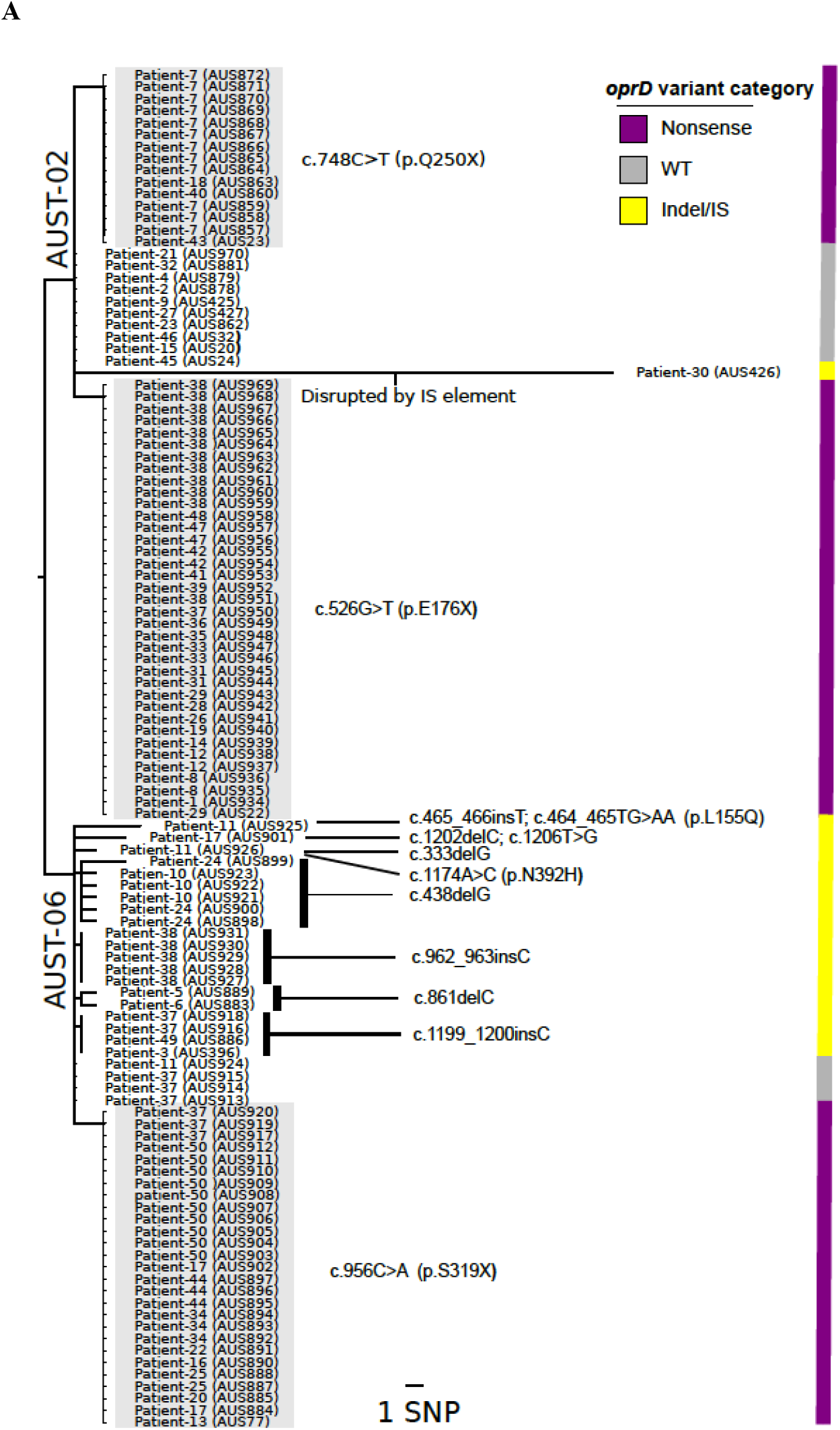
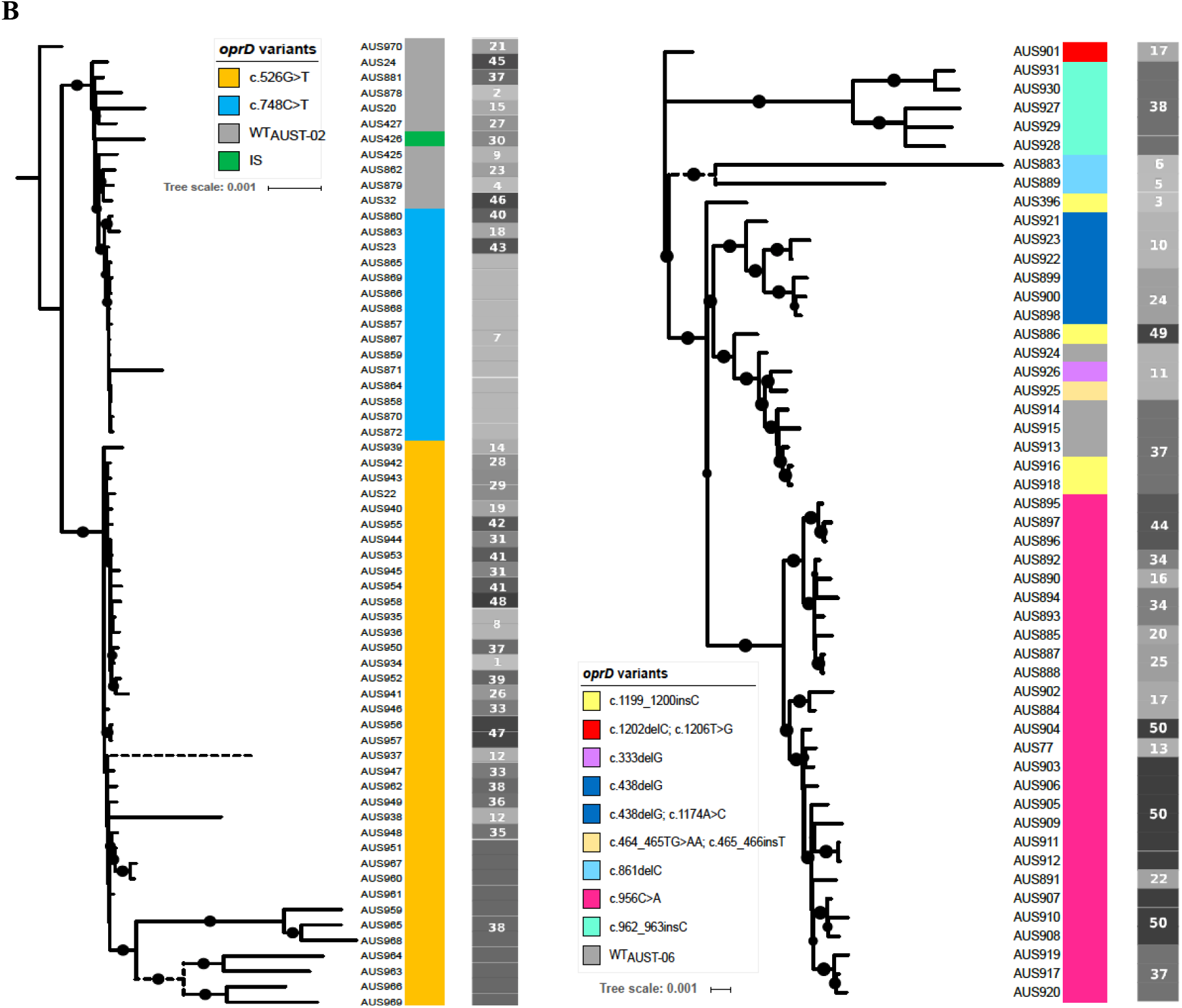
**A** Phylogenetic relationship of AUST-02 and AUST-06 *oprD* variants. Amino acid substitutions shown in parentheses. The Neighbour-Joining algorithm implemented in Jalview was used to visualise relationships between the different variants. The coloured bars on the right-hand side indicate the *oprD* variant categories used to compare AMR results (purple, target SNP i.e. 1-bp substitution leading to nonsense mutation; grey, wild-type sequence; yellow, indel mutation including disruption by IS). **B** Recombination-filtered, Maximum-Likelihood core SNP phylogeny of AUST-02 (left) and AUST-06 (right) isolates encoding the described *oprD* variants. Each variant is given a unique colour (note that one of five isolates with the *oprD* c.438delG variant had acquired an additional *oprD* missense mutation but are denoted by the same colour) in the first column and the patient number is indicated in the 2^nd^ greyscale column. Branches supported by >80% bootstrap are indicated by black circles, branches shown with dotted lines are not to scale. *Definitions*: IS, insertion sequence; WT, wild-type.

Conspicuous loss-of-function mutations in *oprD* caused by a 1-bp substitution or a 1-bp insertion or deletion (indel) occurred multiple times in the strain collection. Notably, three independent nonsense mutations caused by a 1-bp substitution were identified in 79/114 (69%) isolates from 32/50 (64%) patients; two mutations were exclusive to the AUST-02 genotype (*oprD* c.526G>T or *oprD* c.748C>T) and one exclusive to AUST-06 (*oprD* c.956C>A). These SNPs were predicted to result in a truncated protein sequence *in silico*, with *oprD* c.526G>T (p.E176X), c.748C>T (p.Q250X), and c.956C>A (p.S319X) leading to truncation at amino acids 175, 249 and 318, respectively. All AUST-02 isolates carrying the *oprD* c.526G>T (p.E176X) variant belong to the M3L7 sub-type, described previously (Sherrard, Tai et al., 2017, Tai et al., 2015a, Wee et al., 2018).

Seven independent 1-bp indels resulting in a shift in the reading frame were also identified in a smaller proportion of isolates (*n*=20/114; 18%) from 10/50 (20%) participants; each of these belong to the AUST-06 genotype. Six indels were predicted *in silico* to result in a truncated protein sequence, with the remaining indel extending the protein length by 31 amino acids to 472 amino acids.

Only two isolates had acquired additional missense mutations AUS925 (*oprD* c.464_465TG>AA, p.L155Q) and AUS899 (*oprD* c.1174A>C, p.N392H), which were each predicted computationally to affect protein function (Choi, Sims et al., 2012). One isolate (AUS901) had an additional synonymous mutation (*oprD* c.1206T>G). Finally, the *oprD* variant of one AUST-02 isolate (AUS426) was disrupted by an insertion sequence (IS) of the IS3 family that is 100% identical to that found in a sequenced plasmid (GenBank: JX469825.1).

Participants with >1 isolate sequenced, were generally infected with a strain that had an identical *oprD* variant (*n*=14/18; 78%). However, four participants (Patient-11, Patient-17, Patient-24 and Patient-37), had multiple *oprD* variants among their sequenced isolates (Fig 1). Based on the core genome phylogeny of AUST-02 and AUST-06 (Fig 1B), nearly all *oprD* variants emerge from monophyletic clades, which suggests that lineages with specific *oprD* variants pre-existed in the population before spreading between patients. However, we also observed in isolates from Patient-11 and Patient-37, a scenario where multiple *oprD* variants had arisen independently from the same lineage (Fig 1B).

### AMR of clinical isolates

There were no statistical differences in MICs of the six anti-pseudomonal antimicrobials tested between the AUST-02 (*n*=63) and AUST-06 (*n*=51) genotypes (Fig S1A).

Given the prevalence of isolates with a nonsense mutation arising from a 1-bp substitution (‘target SNPs’; *n*=79), we decided to focus on characterising their AMR patterns compared to isolates with other (‘indel’) *oprD* mutations (*n*=21; frameshift caused by 1-bp indel or disruption by IS) or the denoted wild-type variant (*n*=14). MIC comparisons are shown in Fig S1B. Statistically significant differences in MICs were apparent for all antimicrobials except colistin to which 99% of isolates were susceptible (Table I). Isolates with a variant containing a target SNP (100%) or an indel (95%) were highly resistant to imipenem (MIC_50_: >32 mg/L) whereas only one isolate with the wild-type variant (MIC_50_: 1.5 mg/L) was resistant (Table I). Those with a target SNP also demonstrated greater resistance and had statistically higher MICs for meropenem (all *p*<0.001), ceftazidime (*p*=0.04 or *p*<0.001) and ticarcillin-clavulanic acid (all *p*<0.001) relative to isolates with other variants (Table I). Isolates with a target SNP had higher MICs for tobramycin (*p*=0.006) compared to isolates with an indel.

**Table I.** *In vitro* antimicrobial susceptibilities of isolates stratified by *oprD* variant category.

| *oprD* variant category<br>and antimicrobial agent* | MIC (mg/L) |  |  | Percentage of isolates with<br>indicated susceptibility |  |  |
| --- | --- | --- | --- | --- | --- | --- |
|  | Range | 50% | 90% | S | I | R |
| *Target SNP (n=79)* |  |  |  |  |  |  |
| Imipenem | 12 to >32 | >32 | >32 | 0 | 0 | 100 |
| Meropenem | 3 to >32 | >32 | >32 | 0 | 2.5 | 97.5 |
| Ticarcillin-clavulanate | 2 to >256 | >256 | >256 | 8.9 | 2.5 | 88.6 |
| Ceftazidime | 1 to >256 | >256 | >256 | 17.7 | 1.3 | 81.0 |
| Tobramycin | 1 to >256 | 6 | >256 | 40.5 | 29.1 | 30.4 |
| Colistin | 0.125-1.5 | 0.5 | 1 | 100 | 0 | 0 |
| *Indel (n=21)* |  |  |  |  |  |  |
| Imipenem | 12 to >32 | >32 | >32 | 4.8 | 0 | 95.2 |
| Meropenem | 0.75 to >32 | >32 | >32 | 14.3 | 9.5 | 76.2 |
| Ticarcillin-clavulanate | 0.75 to >256 | >256 | >256 | 33.3 | 0 | 66.7 |
| Ceftazidime | 0.25 to >256 | 4 | >256 | 57.1 | 4.8 | 38.1 |
| Tobramycin | 0.5 to >256 | 4 | 64 | 66.7 | 19.0 | 14.3 |
| Colistin | 0.125-3 | 0.75 | 2 | 95.2 | 0 | 4.8 |
| *Wild-type (n=14)* |  |  |  |  |  |  |
| Imipenem | 0.125-6 | 1.5 | 3 | 78.6 | 14.3 | 7.1 |
| Meropenem | 0.012-2 | 0.125 | 1.5 | 100 | 0 | 0 |
| Ticarcillin-clavulanate | 0.125 to >256 | 4 | >256 | 71.4 | 7.2 | 21.4 |
| Ceftazidime | 0.064-16 | 1 | 16 | 85.7 | 14.3 | 0 |
| Tobramycin | 0.25 to >256 | 4 | >256 | 50.0 | 14.3 | 35.7 |
| Colistin | 0.125-2 | 0.5 | 2 | 100 | 0 | 0 |

S, susceptible; I, intermediate; R, resistant
\**oprD* variant categories: Target SNP, one of three 1-bp substitutions leading to a nonsense mutation; indel, 1- bp insertion or deletion leading to a shift in the reading frame or disruption by an insertion sequence; wild-type, variant that is closest to the base of each *oprD* clade in the phylogeny (Fig 1A).

Finally, to indicate if clinical isolates with target SNPs (*oprD* c.526G>T, p.E176X; *oprD* c.748C>T, p.Q250X; *oprD* c.956C>A, p.S319X) had similar AMR profiles, their MICs were compared. Although MICs did not differ for the carbapenems, they varied statistically for the remaining four antimicrobials tested (Fig S1C).

### Allelic replacement of *oprD* in PAO1

Next, we investigated the specific contribution that each target SNP had on carbapenem resistance profiles using PAO1, which is susceptible to imipenem (MIC 1 mg/L) and meropenem (MIC 0.38 mg/L).

Introduction of the observed 1-bp substitutions into *oprD* of PAO1 *via* allelic replacement led to imipenem resistance for all isogenic mutants (PAO1-OprD^E176X^, 32 mg/L; PAO1-OprD^Q250X^, 24 mg/L; PAO1-OprD^S319X^, 32 mg/L) and increased the meropenem MIC by ∼2-3 doubling dilutions (PAO1-OprD^E176X^, 3 mg/L; PAO1-OprD^Q250X^, 2 mg/L; PAO1-OprD^S319X^, 1.5 mg/L).

The meropenem MICs are much lower than the MICs seen in the clinical isolates (MIC_50_: >32 mg/L). All complemented mutants had MICs at wild-type PAO1 levels (Table II). Furthermore, there were no apparent differences in the susceptibility of each isogenic mutant to ceftazidime, ticarcillin-clavulanate, tobramycin and colistin compared to the wild-type strain (Table II).

**Table II.**
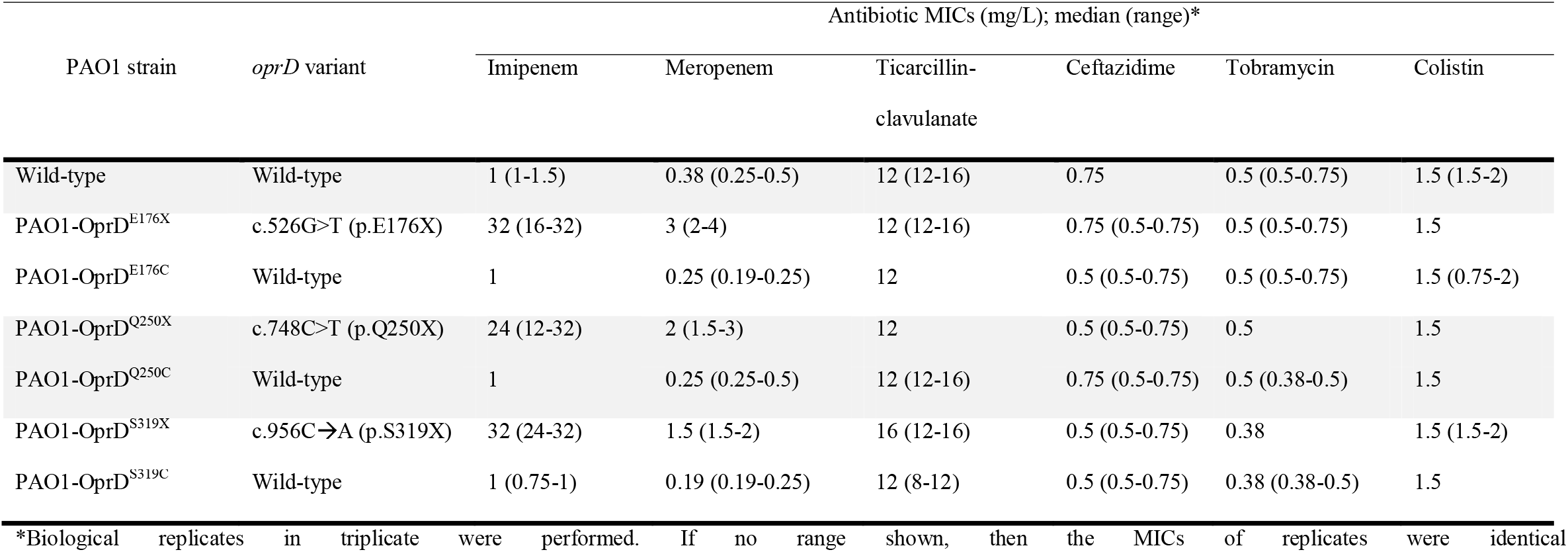
Susceptibility of PAO1 and mutant clones.

| PAO1 strain | *oprD* variant | Antibiotic MICs (mg/L); median (range)* |  |  |  |  |  |
| --- | --- | --- | --- | --- | --- | --- | --- |
|  |  | Imipenem | Meropenem | Ticarcillin-clavulanate | Ceftazidime | Tobramycin | Colistin |
| Wild-type | Wild-type | 1 (1-1.5) | 0.38 (0.25-0.5) | 12 (12-16) | 0.75 | 0.5 (0.5-0.75) | 1.5 (1.5-2) |
| PAO1-OprD<sup>E176X</sup> | c.526G>T (p.E176X) | 32 (16-32) | 3 (2-4) | 12 (12-16) | 0.75 (0.5-0.75) | 0.5 (0.5-0.75) | 1.5 |
| PAO1-OprD<sup>E176C</sup> | Wild-type | 1 | 0.25 (0.19-0.25) | 12 | 0.5 (0.5-0.75) | 0.5 (0.5-0.75) | 1.5 (0.75-2) |
| PAO1-OprD<sup>Q250X</sup> | c.748C>T (p.Q250X) | 24 (12-32) | 2 (1.5-3) | 12 | 0.5 (0.5-0.75) | 0.5 | 1.5 |
| PAO1-OprD<sup>Q250C</sup> | Wild-type | 1 | 0.25 (0.25-0.5) | 12 (12-16) | 0.75 (0.5-0.75) | 0.5 (0.38-0.5) | 1.5 |
| PAO1-OprD<sup>S319X</sup> | c.956C→A (p.S319X) | 32 (24-32) | 1.5 (1.5-2) | 16 (12-16) | 0.5 (0.5-0.75) | 0.38 | 1.5 (1.5-2) |
| PAO1-OprD<sup>S319C</sup> | Wild-type | 1 (0.75-1) | 0.19 (0.19-0.25) | 12 (8-12) | 0.5 (0.5-0.75) | 0.38 (0.38-0.5) | 1.5 |

\*Biological replicates in triplicate were performed. If no range shown, then the MICs of replicates were identical

It was predicted *in silico* that each of the target SNPs would lead to a truncated protein of between 175 and 318 amino acids. A global data-dependent liquid chromatography-mass spectrometry analysis found no evidence of OprD protein in the PAO1-OprD^E176X^, PAO1-OprD^Q250X^, and PAO1-OprD^S319X^ isogenic mutants. In contrast, peptides spanning both N- and C-terminal regions of the OprD protein were detected in PAO1 and all of the complemented mutants. Targeted mass spectrometry using parallel reaction monitoring (PRM) analysis confirmed these results using tryptic peptides that were unique to OprD when compared to the background proteome of PAO1. Finally, *in vitro* growth was not affected by the introduction of the target *oprD* SNPs into the chromosome of PAO1 (Fig S2).

### Clinical course

Variant-specific PCR indicated that 434/890 (49%) isolates from 149 participants had one of the target SNPs in *oprD* (Fig 2).

**Fig 2.**
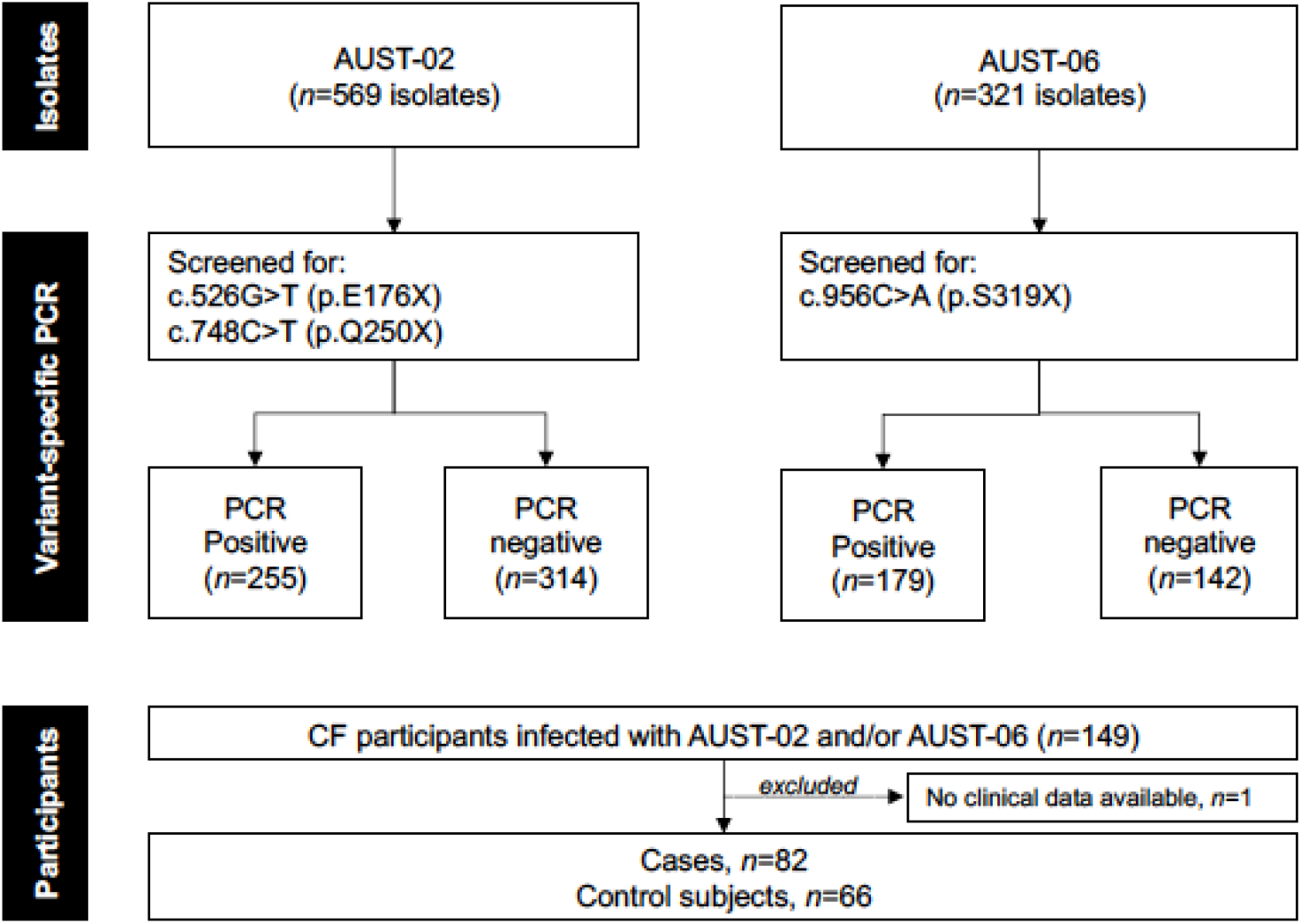
Scheme showing how AUST-02 and AUST-06 isolates were screened for the target *oprD* SNPs. Participants were subsequently stratified according to PCR results: cases, positive for ≥1 *Pseudomonas aeruginosa* isolate with a target SNP; control subjects, isolates PCR negative (i.e. infected with isolates with non-target *oprD* variants).

Overall, 83/149 (56%) participants were infected with ≥1 isolate carrying a target SNP in *oprD* (Fig 2); whereas the remaining participants (control subjects) had infection with isolates with other *oprD* variants. Specifically, 101/569 (18%; 22 participants) and 154/569 (27%; 27 participants) AUST-02 isolates tested positive for the *oprD* c.526G>T (p.E176X) and *oprD* c.748C>T (p.Q250X) variants, respectively. All AUST-02 isolates positive for the *oprD* c.526G>T (p.E176X) variant were confirmed as belonging to the M3L7 sub-type by PCR. More than half (*n*=179/321, 56%; 41 participants) of AUST-06 isolates tested positive for the *oprD* c.956C>A (p.S319X) variant. Seven (5%) participants with >1 isolate had multiple target variants detected in concurrent or longitudinal isolates.

Cases (*n*=82, one case lacked clinical data and was excluded from the analysis) and control (*n*=66) subjects were similar with respect to age, sex, CFTR function, chronic/type of shared strain infection status and co-morbidities. The odds of being a case was reduced with higher lung function (OR, 0.97; 95% CI, 0.96-0.99; *p*<0.001) and increased with a greater number of hospital days (OR, 1.01, 95% CI, 1.00-1.03; *p*=0.010). Although not statistically significant, the odds of being a case was reduced with increasing BMI (OR, 0.92, 95% CI, 0.83-1.02; *p*=0.11). Amongst long-term treatments prescribed, inhaled colistin significantly increased the odds of being a case (OR, 2.48, 95% CI, 1.22-5.04; *p*=0.010) compared to non-inhalation as did tobramycin although it was not significant (OR, 2.13, 95% CI, 0.84-5.43; *p*=0.11) (Table III). After controlling for all these factors in a multivariable logistic regression, only lung function remained statistically significant.

**Table III.** Participant characteristics and their association with being a case.

| Characteristic | Number patients | Cases (n=82)<sup>‡</sup> | Control subjects (n=66)<sup>‡</sup> | Odds ratio (95% CI)<sup>†</sup> | p |
| --- | --- | --- | --- | --- | --- |
| Female; n (%) | 148 | 37 (45.1) | 28 (42.4) | 1.07 (0.54-2.14) | 0.80 |
| *CFTR* function; n (%) | 148 |  |  |  |  |
| - Minimal |  | 70 (85.4) | 57 (86.4) | 1.03 (0.34-3.06) | 1.00 |
| - Residual |  | 4 (4.9) | 2 (3.0) | 0.68 (0.22-2.16) | 0.60 |
| - Non-classified |  | 8 (9.7) | 7 (10.6) | Reference group |  |
| FEV<sub>1</sub> % predicted, mean (sd) | 144 | 49.0 (21.8) | 64.2 (26.1) | **0.97 (0.96-0.99)** | **<0.001** |
| BMI, median (IQR) | 148 | 19.4 (17.2-22.6) | 21.4 (19.3-23.7) | **0.92 (0.83-1.02)** | **0.11** |
| Pancreatic insufficiency; n (%) | 148 | 78 (95.1) | 65 (98.5) | 0.29 (0.03-2.82) | 0.29 |
| *Cystic fibrosis-related diabetes*; n (%) | 148 |  |  |  |  |
| - D+FH |  | 14 (17.1) | 12 (18.2) | 0.92 (0.38-2.22) | 0.84 |
| - IGT |  | 8 (9.7) | 5 (7.6) | 1.25 (0.37-4.17) | 0.72 |
| - Normal GT |  | 60 (73.2) | 49 (74.2) | Reference group |  |
| Liver disease; n (%) | 148 | 9 (11.0) | 7 (10.6) | 1.06 (0.36-3.10) | 0.90 |
| Oral azithromycin; n (%) | 147 | 73 (90.1) | 54 (81.2) | 1.98 (0.73-5.35) | 0.18 |
| Inhaled tobramycin; n (%) | 147 | 71 (86.6) | 51 (77.3) | **2.13 (0.84-5.43)** | **0.11** |
| Inhaled colistin; n (%) | 147 | 46 (56.8) | 24 (36.4) | **2.48 (1.22-5.04)** | **0.010** |
| Inhaled dornase alfa; n (%) | 147 | 60 (74.1) | 43 (65.2) | 1.51 (0.72-3.16) | 0.27 |
| Number of days in hospital, median (IQR) | 148 | 26.5 (10.8-51.8) | 8.5 (0.0-16.8) | **1.01 (1.00-1.03)** | **0.010** |

*Definitions:* D + FH, diabetes with fasting hyperglycaemia; IGT, impaired glucose tolerance; Normal GT, normal glucose tolerance; FEV<sub>1</sub> % predicted, forced expiratory volume in the first second percent predicted; BMI, body mass index; CFTR, cystic fibrosis transmembrane conductance regulator (*CFTR* function: residual function, harbouring ≥1 allele with Class IV-V mutations; minimal function, harbouring two alleles with Class I-III mutations; non-classified, harbouring two alleles with mutations of unknown function).
<sup>‡</sup>Cases, tested positive for ≥1 isolate with a target *oprD* SNP (one case lacked clinical data and was excluded from the analysis); control subjects, tested negative for isolates with a target *oprD* SNP. Data collected relate to the date of first detection of AUST-02 and/or AUST-06 with a target SNP or to the date of first detection of AUST-02 and/or AUST-06 if always tested negative.
<sup>†</sup>All odds ratios presented are adjusted for age and shared strain infection (AUST-02, AUST-06 or mixed) due to their expected effects on odds of testing positive for an isolate with a target SNP in *oprD*; however, these potential confounders were never statistically significant in any of the models. Odds of being a case compared to being a control.
**Bolded** text: variables included in the multivariable logistic regression

Furthermore, 45/148 (30%; one participant lacked clinical data and was excluded from the analysis) participants were deceased (*n*=15) or had received a lung transplant (*n*=30) by the censor date (Table SI). The type of shared strain infection was not found to be predictive of transplant or death (*p*=0.77). However, being a case had a higher risk of transplant or death (HR, 2.44; 95% CI, 1.27-4.68) compared to being a control subject.

## DISCUSSION

The emergence of AMR in bacterial pathogens is a foremost concern for clinical management of people with CF, especially as longevity is increasing, and complex antimicrobial treatment regimens are a mainstay of routine care (Waters, Kidd et al., 2019). This study focused on carbapenems, which are fundamental antimicrobials used intravenously to treat pulmonary exacerbations but are often reserved for those with advanced disease or where there is a suboptimal clinical response to other ß-lactam antibiotics. Therefore, they are a critical class of antimicrobials to manage the most vulnerable patients. Nevertheless, reduced susceptibility of *P. aeruginosa* to carbapenems is reported, including in our Queensland CF population (Smith, Ramsay et al., 2016, Tai, Kidd et al., 2015b); but the underlying resistance mechanisms and the potential clinical implications of their identification are less clear.

In our initial work, we performed a targeted analysis of the *oprD* locus of two major Australian shared strains. We found that both strains frequently carry *oprD* variants, supporting previous studies that identified this gene as a common target for mutation during persistent infection of the CF airways (Greipel et al., 2016, Marvig et al., 2015b). The *oprD* variants identified were not shared by AUST-02 and AUST-06, providing no current evidence of DNA transfer of a variant between the genotypes, even though some patients may be concurrently infected with both strains (Tai, Sherrard et al., 2017). Furthermore, rather than identifying many novel *oprD* variants that were generally exclusive to individual patient isolates, we repeatedly identified three specific variants in isolates from multiple patients, which were characteristic of monophyletic strain sub-lineages (Fig 1B). Each variant was truncated due to a nonsense mutation caused by a SNP. We hypothesise that these SNPs became fixed in the sub-lineages following their divergence from all other AUST-02/AUST-06 strains and these sub-lineages were subsequently disseminated between patients, possibly from patient socialisation or via the aerosol mode of transmission (Knibbs, Johnson et al., 2014, Ojeniyi, Frederiksen et al., 2000). This is supported by finding that all AUST-02 isolates positive by PCR for the *oprD* c.526G>T (p.E176X) variant were confirmed as belonging to the recently characterised AUST-02 sub-lineage, M3L7, and provides further evidence that this mutation is exclusive to the lineage (Wee et al., 2018). In addition, introduction of the various SNPs of interest into *oprD* of PAO1 had no impact on *in vitro* growth suggesting that these mutations may be acquired without a cost to fitness. Acquiring mutations without a fitness cost may further substantiate the likelihood of clinical strains carrying these *oprD* variants successfully spreading and establishing new infections in the CF patient population (MacLean & San Millan, 2019).

Interestingly, we also observed two cases where a single AUST-06 lineage acquired two or three different *oprD* variants within a patient (Fig 1B). It is unclear whether these variants were already present in the AUST-06 population before the patient was infected or whether each variant arose independently during the course of infection.

It is reported that reduced expression or inactivation of OprD constitutes a major resistance mechanism to carbapenems (Courtois et al., 2018, Li, Luo et al., 2012, Llanes, Pourcel et al., 2013) and in keeping with this, our analysis revealed that 99% of imipenem-resistant isolates harboured *oprD* with either a nonsense or indel mutation or on one occasion, disruption by an insertion sequence (MIC_50_: >32mg/L). Conversely, a full-length coding sequence was associated with much lower MICs (MIC_50_: 1.5mg/L). One clear discrepancy was a single isolate (AUS396) carrying a 1-bp insertion in *oprD* between nucleotides at positions 1199 and 1200, which was susceptible to imipenem (MIC: 2mg/L). This frameshift mutation was predicted to extend the length of the protein and indeed an earlier study found that isolates carrying additional amino acids at the C-terminus of OprD remained susceptible to carbapenems (Ocampo-Sosa, Cabot et al., 2012). However, three other isolates (AUS886, AUS916, AUS918) carrying the same 1-bp insertion as AUS396 showed resistance to imipenem (MICs: >32mg/L). This highlights the complexity of deciphering the development of AMR and the need for studies characterising the impact of individual and combinations of chromosomally acquired mutations.

The impact of the various mutations on porin integrity is influenced by their position in the protein sequence (Epp, Kohler et al., 2001, Huang, Jeanteur et al., 1995, Ocampo-Sosa et al., 2012). We determined the effect of the three most common *oprD* SNPs in our strain collection using site-directed mutagenesis in the reference strain, PAO1. Our results showed imipenem resistance and an apparent loss of the porin for all three truncations (p.E176X, p.Q250X, p.S319X). Although 97% of clinical isolates with the target SNPs also had a meropenem MIC >32mg/L, we found that in our isogenic mutants the target SNPs increased the meropenem MIC only by several fold to a maximum of 3 mg/L. It is recognised that mutations in *oprD* alone do not cause meropenem resistance (Strateva & Yordanov, 2009). The target *oprD* variants are likely to contribute to reduced susceptibility to meropenem in the clinical isolates and clinical resistance may be a polygenic trait. More genetically diverse collections of isolates with meropenem resistance are needed to identify the genes that are significantly associated with this phenotype. Additionally, the SNPs did not impact the MICs of other β-lactam antibiotics, tobramycin or colistin confirming that these mutations specifically affect the activity of carbapenem antibiotics.

Based on our prior genome sequencing studies of the M3L7 sub-lineage (Sherrard et al., 2017, Wee et al., 2018), it is likely that the individual clinical isolates included in the current study harbor a multitude of concurrent mutational mechanisms to achieve the high-level resistance observed to carbapenems and the other antimicrobials studied. Carbapenem resistance can arise from the production of antibiotic-inactivating carbapenemases; but recent studies failed to detect acquired carbapenemase encoding genes in CF *P. aeruginosa* isolates suggesting that, to date, this mechanism is uncommon in CF (Chalhoub, Saenz et al., 2016, Courtois et al., 2018, Mustafa, Chalhoub et al., 2016, Tai et al., 2015b). Upregulation of multi-drug efflux pumps and de-repression of the cephalosporinase, AmpC, harbouring spectrum-extending mutations may also contribute to increased MICs (Rodriguez-Martinez, Poirel et al., 2009, Strateva & Yordanov, 2009). We previously identified mutations in various efflux pump regulators (e.g. *mexZ, nalC, nalD*) and pump proteins (e.g. *mexX, mexY*) and *ampC* which might also contribute to carbapenem resistance in M3L7 (Sherrard et al., 2017). It was proposed that inactivation of OprD could be the primary resistance mechanism induced by carbapenem selective pressure with further mechanisms acquired thereafter (Li et al., 2012).

The AUST-02 and AUST-06 genotypes, have been associated with increased treatment requirements compared to unique *P. aeruginosa* genotypes (Kidd et al., 2013). Prior studies have also identified that adaptive mutations in specific loci, including in *mucA* or related genes that lead to mucoidy or loss-of-function mutations in the transcriptional regulator of quorum sensing *lasR* were associated with accelerated decline in lung function or disease progression (Hoffman et al., 2009, Li et al., 2005). In our current work, we identified a subset of people (>50%) infected with AUST-02/AUST-06 with poorer clinical outcomes based on detection of specific *oprD* variants. This finding extends our previous work that identified a small group of adults infected with M3L7 who had a greater 3-year risk of lung transplantation/death (Tai et al., 2015a) and further highlights the heterogenous clinical course of people despite infection with clonally-related strains. When we investigated the characteristics of people who had acquired strains carrying the target *oprD* SNPs in our multivariable model, lower forced expiratory volume in one-second %predicted was potentially important. As we are unable to precisely determine when each person first became infected with these strains, it is unknown whether they contribute to lung function decline or alternatively, if certain individuals were at greater risk of their acquisition e.g. from spending more time in hospital; a statistically significant variable in the univariable analysis. Moreover, we found a relationship between people infected with a strain carrying a target *oprD* SNP and subsequent increased hazard rate of lung transplantation or death; but this also does not establish causality. Other studies of *P. aeruginosa* blood stream infections have reported that loss of OprD is associated with mortality (Fluit, Rentenaar et al., 2019, Yoon, Kim et al., 2019).

A recent study showed that *P. aeruginosa* PA14 transposon *oprD* mutants were carbapenem resistant, and also had enhanced serum resistance and survival in a low pH environment and cytotoxicity against phagocytes in a murine model, which possibly occurred *via* alteration of expression of other genes (Skurnik et al., 2013). Therefore, further work is required to characterise if the target *oprD* SNPs identified in our study also lead to the emergence of more virulent CF sub-lineages with enhanced survival in the acidified airway surface liquid that is characteristic of the CF lungs (Pezzulo, Tang et al., 2012).

Limitations of this study include the small number of isolates that were available per person, especially as infection with multiple strain types and co-existence of divergent strain sub-lineages is recognised (Diaz Caballero et al., 2015, Tai et al., 2017, Williams et al., 2015). Therefore, we may have underestimated the proportion of people infected with strains carrying the target variants. In addition, the variant-specific PCRs were specific to the SNPs of interest and therefore, only have a potential prognostic value to patient populations where AUST-02 and AUST-06 genotypes have been identified. Furthermore, it is likely that the proportion of isolates, which were negative for the target SNPs, will harbour other loss-of-function *oprD* mutations. If these mutations have similar features to the target SNPs is unclear. It is also unknown if the *oprD* SNPs identified have a therapeutic value in predicting response to pulmonary exacerbation treatment regimens containing carbapenems. Currently, *in vitro* AMR and *in vivo* response to treatment are not well correlated in chronic *P. aeruginosa* infection (Somayaji, Parkins et al., 2019). Finally, we were unable to exchange the *oprD* variant carrying a nonsense mutation in AUST-02 or AUST-06 clinical isolates with that of the wild-type variant, as the clinical isolates were insensitive to SacB-mediated sucrose toxicity used for counter selection.

## CONCLUSIONS

We found that nucleotide substitutions and indels were commonly encountered in the *oprD* gene of two prevalent shared strains in an Australian CF population and included three independently acquired nonsense mutations fixed in different sub-lineages. These nonsense mutations could largely explain imipenem resistance amongst the corresponding CF isolates and may be a marker of worse patient outcomes. Studying genes under strong selection pressure, such as *oprD*, in detail may help improve our understanding of chronic-stage adaptions that contribute to strain transmissibility and greater adverse patient outcomes observed in some people.

## MATERIALS AND METHODS

### Clinical isolates

To account for potential diversity in AMR *in vitro* and *oprD* sequences, two shared strains, AUST-02 and AUST-06, were selected for this study. All isolates were obtained by prior surveillance studies between 2001-2014 (Kidd et al., 2013, O’Carroll, Syrmis et al., 2004, Ramsay, Bell et al., 2016, Sherrard et al., 2017) from single colony sputum cultures and were identified as AUST-02 or AUST-06 *via* ERIC-PCR and/or iPLEX20SNP genotyping (Kidd et al., 2013, Kidd, Ramsay et al., 2015, O’Carroll et al., 2004, Ramsay et al., 2016, Sherrard et al., 2017, Syrmis, Kidd et al., 2014). Isolates are held in the QIMR Berghofer Pseudomonas Biobank.

### Ethics approval

Ethics approval was granted under HREC/07/QRCH/9 and HREC/13/QPCH/127 by The Prince Charles Hospital (TPCH) Human and Research Ethics Committee. All participants provided written, informed consent/assent.

### Whole genome analysis

Genomic DNA from 63 AUST-02 isolates (including 37 isolates of the AUST-02 strain sub-type, M3L7; PRJEB14781, PRJEB14771, PRJEB21755) and 51 AUST-06 isolates (PRJEB35026, PRJNA325248) were utilised (Freschi et al., 2019, Sherrard et al., 2017, Tai et al., 2015a, Wee et al., 2018). These isolates originated from the sputum of 50 participants (10 aged <18-years) attending three CF centres in Brisbane (see Table SII for further details). One isolate was sequenced from 32 participants with a median of three (range, 2-17) isolates sequenced from the remaining 18 participants. Two participants had both strains identified contemporaneously (Patient-37 & Patient-38). Whole genome sequences were evaluated for contamination and quality filtered using Nesoni clip (v0.128) (https://github.com/Victorian-Bioinformatics-Consortium/nesoni). Genomes were assembled using Velvet (v1.2.10) (Zerbino & Birney, 2008) and VelvetOptimiser (v2.2.5) (https://github.com/tseemann/VelvetOptimiser). Contigs were reordered with PAO1 as the reference using Mauve (v2.4.0) (Darling, Mau et al., 2010). Assemblies were annotated with Prokka (v1.10) with the PAO1 genome as the primary source of annotation (Seemann, 2014).

Due to the absence of a closely related complete reference genome (less than 20,000 single nucleotide polymorphisms [SNPs]) to AUST-02 and AUST-06 lineages, we first used Parsnp to align the AUST-02 and AUST-06 isolates to PAO1 to determine the approximate position of the root of each lineage (Treangen, Ondov et al., 2014). A multiple genome alignment was then calculated with only genomes from each shared strain. Alignments were filtered for recombination using Gubbins (v2.3.1) and Maximum-Likelihood phylogenies were constructed with IQTREE (v1.6.6) with 1000 bootstrap replicates (Croucher, Page et al., 2014, Nguyen, Schmidt et al., 2015).

### Identification of *oprD* variants

Gene sequences of *oprD* from the 63 AUST-02 and 51 AUST-06 isolates were then obtained from the whole genome sequences. Sequence similarity to *oprD* from PAO1 (PA0958; 1332-base pairs [bp]) was determined using blastn requiring ≥50% of the query sequence to be present. The AUST-02 and AUST-06 *oprD* variants were then aligned using TranslatorX and Mafft (Abascal, Zardoya et al., 2010, Katoh & Standley, 2013). Relationships between the different *oprD* variants were visualised using the Neighbour-Joining algorithm implemented in Jalview (v2.10.4b1) (Waterhouse, Procter et al., 2009).

### Etest® susceptibility testing

Isolates were inoculated onto Mueller-Hinton Agar and minimum inhibitory concentrations (MICs) were obtained by Etest^®^ (BioMérieux) according to the manufacturer’s instructions. Isolates were categorised as susceptible, intermediate or resistant (CLSI, 2018). The *P. aeruginosa* ATCC 27853 strain was used for quality control.

### Site-directed mutagenesis

Bacterial strains and plasmids are described in Table SIII. Strains were cultured in Luria-Bertani broth at 37°C with shaking at 200rpm and supplemented with antibiotics when appropriate: kanamycin 50µg/mL or streptomycin 50µg/mL or 2000µg/mL (Astral Scientific). Polymerase chain reaction (PCR) assays were performed in an Applied Biosystems Veriti Thermal cycler with oligonucleotide primer sequences shown in Table SIV.

Mutants of *P. aeruginosa* PAO1 (Klockgether, Munder et al., 2010), were constructed by site-directed mutagenesis of chromosomally encoded *oprD*, performed essentially as described by Muhl and Filloux (Muhl & Filloux, 2014); specific details and minor variations are outlined in the supporting information. Plasmid constructs with the SNPs of interest (*oprD* c.526G>T, p.E176X; *oprD* c.748C>T, p.Q250X; *oprD* c.956C>A, p.S319X) were constructed using plasmid pMRS101 (Sarker & Cornelis, 1997). The pMRS101 constructs were mobilised from *Escherichia coli* CC118 λ pir into PAO1 using a triparental mating with *E. coli* DH5α harbouring the helper plasmid pRK2013 (Ditta, Stanfield et al., 1980) according to the method of Sana *et al* (Sana, Laubier et al., 2014). A single homologous recombination event was selected using streptomycin and a second homologous recombination event was counter selected with 5% sucrose was performed using the method described by Muhl and Filloux (Muhl & Filloux, 2014). The disc diffusion method (Thermo Fisher Scientific) was employed to detect resistance to imipenem as a screen for potential isogenic mutants. The presence of the SNPs in the genome of putative PAO1 mutants was confirmed by PCR and DNA sequencing. The confirmed isogenic mutants were named: PAO1-OprD^E176X^, PAO1-OprD^Q250X^, and PAO1-OprD^S319X^.

Complementation of the PAO1-OprD^E176X^, PAO1-OprD^Q250X^, and PAO1-OprD^S319X^ mutants was performed using a pMRS101 plasmid construct with the wild-type *oprD* gene encoded and the same methodologies employed to generate the original mutations. Complemented mutants were named: PAO1-OprD^E176C^, PAO1-OprD^Q250C^ and PAO1-OprD^S319C^.

### Proteomic analysis of OprD from *P. aeruginosa* PAO1

A detailed description of all steps is provided in the supporting information. Briefly, C18 reverse phase chromatography was performed on whole protein preparations from the cell pellets of PAO1, each isogenic mutant and its complement. Data-dependent acquisition (DDA) runs were performed to identify sequence coverage of the OprD protein and generate a spectral library and scheduled targeted PRM runs for targeted verification. Protein identification was performed using MaxQuant (v1.6.2.6) (Cox & Mann, 2008, Cox, Neuhauser et al., 2011). The results were visualised using Perseus (Tyanova, Temu et al., 2016). Skyline was used to generate scheduled targeted methods and select precursor-product pairs (transitions) from the DDA dataset (MacLean, Tomazela et al., 2010, Schilling, Rardin et al., 2012).

### Growth curve analysis

As an indicator of *in vitro* fitness, the growth of each isogenic mutant was compared to that of PAO1. 200µL of a culture containing 1.0–1.5 × 10^6^ colony-forming units/mL in Luria-Bertani Broth was transferred to a 96-well microtitre plate in triplicate and incubated with shaking at 37°C for 20-hours. Growth was monitored (OD_620nm_) using a Biotek Powerwave plate reader.

### Variant-specific PCR

Variant-specific PCR assays were designed to rapidly screen a collection of AUST-02 (*n*=569) and AUST-06 (*n*=321) isolates for target *oprD* SNPs. These isolates originated from sputum of 149 participants (median [range], 6 [1-19] isolates/person). Primer sequences are described in Table SV. Each reaction mix contained 10µL Platinum SYBR Green qPCR superMix-UDG (Invitrogen), 1µL each of 10µM sense and anti-sense primer pairs (Sigma-Aldrich), 2µL of isolate DNA (Anuj, Whiley et al., 2009) and nuclease-free water (Qiagen) made up to a total volume of 20µL. The PCR amplification was performed on a Rotor-Gene Q instrument (QIAGEN Pty Ltd) as described in the supporting information.

### Clinical characteristics

Data were retrieved from medical records, the referring CF centre and/or the Australian CF Data Registry. Age, gender, height, weight, spirometry with percent predicted calculated using the Global Lung Function Initiative equations (Quanjer, Hall et al., 2012), co-morbidities, long-term treatments and infection status with *P. aeruginosa* (Lee, Brownlee et al., 2003, Ramsay, Sandhu et al., 2017) were obtained. Cystic fibrosis transmembrane conductance regulator (CFTR) function was recorded from the genotype as residual, minimal or non-classified (Green, McDougal et al., 2010). Data on patients’ hospitalisations in the previous year were collected. The date of death or lung transplantation was recorded, where applicable.

### Data analyses

Statistical analyses were generated using IBM SPSS v22. MICs were compared using a Mann-Whitney *U* test or a Kruskal-Wallis test with the Bonferroni adjustment applied. Categorical data were compared using Pearson’s Chi-squared test. Based on the outcome of the variant-specific PCRs, participants were categorised as either cases (PCR positive for a target *oprD* SNP) or control subjects (PCR negative) and clinical characteristics of each group were described using descriptive statistics. For cases, data relate to the date of first detection of a strain that tested positive for a target SNP. For controls, data relate to the date of first detection of AUST-02 or AUST-06. Participants (*n*=9) with only one isolate were reported according to that PCR result. Potential predictors of being a case were identified by univariate analysis as having p≤0.15 and examined in a multivariable logistic regression model using backwards stepwise selection; variables expected to affect risk (age and type of shared strain infection) were retained to control for any potential confounding effects. Prior to their inclusion variables were checked for collinearity. Adjusted odds and 95% confidence intervals are reported. Cox regression was used to assess the association between being a case with time to lung transplantation or death. For patients who had not had a transplant and were still alive, time was censored on the 31^st^ August 2017. The Cox regression model also included age, gender and the type of shared strain infection. The hazard ratio and 95% confidence interval are reported.

## Data Availability

Genome sequence data available at the European Nucleotide Archive under studies PRJEB14781, PRJEB14771, PRJEB21755, PRJEB35026 and PRJNA325248

## ACKNOWLEDGEMENTS

We thank the Proteomics Facility of QIMR Berghofer Medical Research Institute, and the advice of Emma Norris, Dr. Madeleine Headlam, and Dr. Harsha Gowda relating to the proteomics aspect of this work is greatly appreciated. We also thank Chrissie Schot and Samantha Webster (QIMR Berghofer Medical Research Institute) for their contribution to the site-directed mutagenesis and variant-specific PCRs aspects of this study, respectively.

## AUTHOR CONTRIBUTIONS

LJS, BAW, TJK and SCB conceived the study and wrote the first draft of the manuscript. LJS, BAW, CD, KAR, KAD, EB, CEW, KG, HES, DMW, SAB, TJK and SCB designed/performed the experiments, performed analysis, interpreted the results and/or contributed data for this work. All authors contributed to and approved the final version of the manuscript.

## THE PAPER EXPLAINED

### Problem

*Pseudomonas aeruginosa* is the most important pathogen chronically infecting the lungs of people with cystic fibrosis (CF) and cross-infection of strains (shared strains) can occur. *P. aeruginosa* is known to develop antimicrobial resistance (AMR) including to carbapenem antibiotics, which are often reserved for people with the most advanced disease. Despite reports of reduced susceptibility of CF *P. aeruginosa* to carbapenems, the underlying resistance mechanisms are less clear. Studies have also shown that as *P. aeruginosa* adapts to persist in the CF lungs during long-term antibiotic treatment, certain genes are more prone to mutation than others. However, there is limited information on whether mutations in AMR-related genes (e.g. *oprD*) are characteristic of particular shared strain genotypes or whether novel variants are generally observed in individual patient isolates. Importantly, there is a paucity of data on whether the chromosomal mutations observed are of any clinical relevance.

## Results

In this study, we performed a detailed analysis of the *oprD* gene, which is hypothesised to play an important role in adaptation of *P. aeruginosa* to the CF lungs, and AMR in two shared strains that are prevalent in our patient population. We identified commonly occurring *oprD* allelic variants in isolates from multiple patients, that were unique to specific shared-strain lineages suggesting that successful *oprD* alleles emerge and persist in these shared strain populations. By constructing and characterising *P. aeruginosa oprD* mutants, we show that these variants are likely to contribute to reduced carbapenem susceptibility without compromising *in vitro* fitness. We also identified a subset of people infected with the shared strains with poorer clinical outcomes, including an increased hazard rate of death or lung transplantation, based on detection of specific *oprD* variants.

### Impact

Our findings emphasise the role of *oprD* mutations in carbapenem resistance in the setting of *P. aeruginosa* lung infections in CF. Further our findings highlight the importance of thorough investigations of genes under strong antimicrobial selection pressure to help improve our understanding of (1) chronic-stage *P. aeruginosa* adaptions that may contribute to strain transmissibility and (2) the greater adverse patient outcomes observed in some people with CF despite infection with clonally-related strains.

